# Individual fixel-based white matter abnormalities in the epilepsies

**DOI:** 10.1101/2023.03.16.23287290

**Authors:** Remika Mito, Mangor Pedersen, Heath Pardoe, Donna Parker, Robert E. Smith, Jillian Cameron, Ingrid E. Scheffer, Samuel F. Berkovic, David N. Vaughan, Graeme D. Jackson

**Affiliations:** Florey Institute of Neuroscience and Mental Health, Heidelberg, Victoria, Australia; Florey Department of Neuroscience and Mental Health, University of Melbourne, Melbourne, Victoria, Australia; Department of Psychology and Neuroscience, Auckland University of Technology (AUT), Auckland, New Zealand; Epilepsy Research Centre, Department of Medicine, University of Melbourne, Austin Health, Heidelberg, Victoria, Australia; Department of Neurology, Austin Health, Heidelberg, Victoria, Australia

## Abstract

Diffusion MRI has provided insight into the widespread structural connectivity changes that characterise the epilepsies. Although syndrome-specific white matter abnormalities have been demonstrated, studies have predominantly relied on statistical comparisons between patient and control groups. For diffusion MRI techniques to be of clinical value, they should be able to detect white matter microstructural changes in individual patients. In this study, we apply an individualised approach to a novel technique known as fixel-based analysis, to examine fibre-tract-specific abnormalities in individuals with epilepsy. We explore the potential clinical value of this individualised fixel-based approach in epilepsy patients with differing syndromic diagnoses. Diffusion MRI data from 90 neurologically healthy control participants and 10 patients with epilepsy (temporal lobe epilepsy, Progressive Myoclonus Epilepsy, Dravet Syndrome, malformations of cortical development) were included in this study. Measures of fibre density and cross-section were extracted for all participants across brain white matter fixels, and mean values computed within select tracts-of-interest. Scanner harmonised and normalised data were then used to compute Z-scores for individual patients with epilepsy. Microstructural white matter abnormalities were observed in distinct patterns in individual patients with epilepsy, both at the tract and fixel level. For patients with specific epilepsy syndromes, the detected white matter abnormalities were largely in line with expected syndrome-specific clinical phenotypes. In patients with lesional epilepsies (e.g., hippocampal sclerosis, periventricular nodular heterotopia, bottom-of-sulcus dysplasia), microstructural abnormalities were concordant with lesion location. This study demonstrates the clinical potential of translating advanced diffusion MRI methodology to individual patient-level use in epilepsy. This technique could be useful both in aiding diagnosis of specific epilepsy syndromes, and in localising structural abnormalities, and is readily amenable to other neurological disorders. We have included code and data for this study, so that individualised white matter changes can be explored robustly in larger cohorts in future work.

## Introduction

Epilepsy is widely considered a network disorder, with hyperexcitability and hypersynchrony within brain networks driving disturbance in brain function^1^. Although the study of brain network disturbances in epilepsy has predominantly focused on abnormal functional connectivity, there is growing interest in changes to the brain’s structural connections, which likely subserve these functional network disruptions. Indeed, both focal and generalised epilepsies have been shown to exhibit widespread abnormalities in white matter fibre pathways, and these structural network disruptions are now understood to be a key characteristic of the epilepsies^1,2^.

Diffusion MRI or diffusion-weighted imaging (DWI) is currently the only tool available to non-invasively probe the *in vivo* white matter architecture of the brain, and has played a crucial role in untangling structural network changes in epilepsy. Diffusion MRI studies have revealed syndrome-specific patterns of white matter alterations, both within predefined cohorts^3–6^, and across large consortia studies^2^. These white matter changes appear clinically meaningful, as they are associated with cognitive function^7,8^ and postsurgical outcome^9,10^. In epilepsy patients with focal brain abnormalities, diffusion MRI measures may also help to localise epileptogenic lesions^11,12^.

Over the past two decades, major technical advancements in diffusion MRI have brought corresponding potential for clinical utility in epilepsy. Yet, in clinical *practice*, use of this technique is limited to a few well-defined applications; for example, to identify and avoid the optic radiation or corticospinal tract when planning surgical interventions in focal epilepsy^13^. This could be attributed to diffusion MRI studies in epilepsy being predominantly focused on group analyses, comparing patient cohorts with control cohorts. These group-level studies are inherently desensitised to individual patient differences, where clinical value is likely to be derived. For advanced neuroimaging techniques to be valuable in clinical practice, they must provide biologically and clinically meaningful information about individuals.

Unfortunately, there are challenges in translating diffusion MRI methods into individual patient use. Firstly, DWI metrics often relate to features of the observed diffusion process rather than direct properties of the underlying biology, such that clinical interpretation is often difficult^14,15^. Secondly, the detection of individual-level changes in neuroimaging-derived measures is hampered by the multiple comparison problem when using classical statistical frameworks. Finally, generalisability of diffusion MRI measures is difficult, given that their values depend on factors such as acquisition site and scanning protocol^16–18^.

In this study, we apply the Fixel-Based Analysis (FBA) framework at the individual patient level, to examine white matter fibre tract-specific changes in individuals with epilepsy. Unlike the more commonly used voxel-averaged measures derived from techniques like diffusion tensor imaging (DTI), FBA involves modelling of multiple fibre orientations within a voxel^19,20^, such that derived measures are both sensitive and specific to individual fibre orientations within a voxel. This provides more biologically interpretable measures of white matter fibre tract changes than can DTI, particularly in crossing-fibre voxels that constitute the majority of brain white matter^21^.

Here, we provide the first (to our knowledge) individualised framework for fixel-based analysis, and examine fibre tract-specific changes in individuals with various causes of epilepsy. We provide means to quantify and visualise individual-level tract changes and incorporate data harmonization approaches into the fixel-based framework, including code and data for further exploration. We demonstrate clinically meaningful changes in individual epilepsy patients, which could be useful in the clinical work-up of syndromic epilepsies, and in localising small epileptogenic lesions.

## Methods

### Participants

Ten individuals with epilepsy were included in this study: 2 subjects with temporal lobe epilepsy and hippocampal sclerosis; 2 subjects with MRI-negative temporal lobe epilepsy; 2 subjects with focal cortical dysplasia (bottom of the sulcus dysplasia); 2 subjects with periventricular nodular heterotopia (1 who had additional polymicrogyria); 1 subject with Dravet’s syndrome; and 1 subject with Progressive Myoclonic Epilepsy. People with epilepsy were recruited through the Austin Health Comprehensive Epilepsy Program, First Seizure clinic, and/or neurology department at Austin Health, and had a confirmed diagnosis of epilepsy. Patients were selected from previous imaging studies where we performed group comparisons between patients with specific epilepsy syndromes or causes and control participants. For this proof-of-principle study, we selected at patients who exhibited clinical signs that were representative of the particular syndrome or cause of epilepsy, rather than patients who showed complex etiology. Clinical data for each patient included in this study is available in Table 1.

**Table 1:** Clinical characteristics of epilepsy patients included in study.

| Patient | Sex | Age range at scan (years) | Epilepsy syndrome or cause | Lesion location | Age at seizure onset (years) |
| --- | --- | --- | --- | --- | --- |
| 1 | M | 26-30 | L TLE-HS | L temporal lobe | 6-10 |
| 2 | F | 41-45 | R TLE-HS | R temporal lobe | 6-10 |
| 3 | M | 16-20 | R TLE-LN | - | 16-20 |
| 4 | F | 26-30 | L TLE-LN | - | 11-15 |
| 5 | M | 26-30 | PME | - | 11-15 |
| 6 | M | 26-30 | Dravet | - | 0-5 |
| 7 | F | 41-45 | PVNH | L lateral ventricles | 6-10 |
| 8 | M | 21-25 | PVNH + PMG | Right ventricular/temporal | 16-20 |
| 9 | M | 16-20 | BOSD | R precentral sulcus | 0-5 |
| 10 | F | 16-20 | BOSD | R postcentral sulcus | 11-15 |

*BOSD: bottom-of-sulcus dysplasia; PME: Progressive Myoclonus Epilepsy; PMG: polymicrogyria; PVNH: periventricular nodular heterotopia; TLE-HS: temporal lobe epilepsy with hippocampal sclerosis; TLE-LN: lesion-negative temporal lobe epilepsy. Age ranges are provided for participants (5-year age range).*

Ninety control participants were included in the normative sample. Control participants were adults (mean age: 36.9 years (±13.3), range: 18 – 64 years) who had no history of seizures, psychiatric illness or significant head injury.

Written informed consent was provided from all participants or their parents/guardians. The study was approved by the Austin Health Human Research Ethics Committee.

### MRI data

All participants underwent an MRI scan at the Florey Institute of Neuroscience and Mental Health. MRI data were acquired at 3T either on a 3T Siemens Skyra or 3T Siemens Trio between 2011 and 2019.

Diffusion-weighted imaging (DWI) data were acquired on the Skyra with a 20-channel head coil receiver, with the following parameters: 60 axial slices, TR/TE = 8400/110 ms, 2.5 mm isotropic voxels, in-plane parallel acceleration factor 2, 64 diffusion-weighted images (b=3000s/mm2) and at least 1 b=0 image. Equivalent DWI were acquired on a Siemens Tim Trio with a 12-channel head coil receiver with the following parameters: 60 axial slices, TR/TE = 8300/110 ms, 2.5 mm isotropic voxels, in-plane parallel acceleration factor 2, 60 diffusion-weighted images (b=3000 s/mm2) and 8 b=0 images. A reverse phase-encoded b=0 image was acquired in all cases to correct for B0 field inhomogeneities.

Isotropic T1-weighted magnetization-prepared acquisition gradient echo (MPRAGE) images were also acquired from all participants with the following parameters: TR/TE = 1900/2.5 ms, inversion time = 900 ms, flip angle = 9°, voxel size = 0.9 mm3, acquisition matrix 256 × 256 × 192. Intracranial volume was computed from T1-weighted images using SPM12^22^.

### Diffusion-weighted image processing

All DWI data were preprocessed and analysed primarily using MRtrix3^23^, with several specific tasks deferred to other software as described below.

DWI data from all participants were preprocessed prior to analysis. Data were first denoised^24^, after which Gibbs ringing artefacts were removed^25^. Susceptibility distortion correction, eddy-current and motion correction were then performed using FSL’s “topup” and “eddy” tools (version 6.0)^26–28^. Bias field correction was then performed based on the mean *b*=0 image using ANTs N4^29^, and DWI images were upsampled to a voxel size of 1.3mm^2^ using cubic b-spline interpolation.

Following these pre-processing steps, fibre orientation distribution functions (ODFs) were computed using Single-Shell 3-Tissue Constrained Spherical Deconvolution (SS3T-CSD)^30^, using group-averaged response functions for white matter (WM), grey matter (GM), and CSF^31,32^. Joint bias field correction and intensity normalization were then performed^33,34^.

A ‘healthy’ population template image was generated using WM ODF images from a subset of 20 healthy control participants selected randomly from the control cohort, using an iterative registration and averaging approach^35^. Spatial correspondence was achieved by registering WM ODF images from all subjects (controls and patients) to this template using ODF-guided non-linear registration^35,36^.

A whole-brain tractogram was generated using probabilistic tractography on the population template image. Twenty million streamlines were generated, after which the SIFT algorithm was applied to filter the tractogram to 2 million streamlines to reduce reconstruction biases^37^.

We then applied the fixel-based analysis (FBA)^20^ framework, where the term ‘fixel’ refers to a specific fibre population within a single image voxel. Different voxels within an image may contain a different number of fixels. These are computed based on segmentation of the WM ODFs^37^ with correspondence established between subject-specific fixels and those of the WM ODF template^20^. A measure of fibre density and cross-section (FDC) was obtained at each white matter fixel in the population template space for all subjects. This FDC metric combines microstructural and morphological information, such that it is sensitive to changes in the density of fibres passing in a particular direction within a given voxel, as well as the changes in the cross-section of fibre bundles that traverse multiple image voxels^20^. Connectivity-based smoothing was performed on fixel-based measures for all participants^38^.

### Tract-level analysis

Major fibre tracts of interest were delineated on the WM ODF template using TractSeg^39^. Tracts of interest were selected for analysis based on having previously exhibited abnormality in group studies comparing epilepsy patients to control cohorts were selected for analysis. These tracts were: the arcuate fasciculus (AF), corpus callosum (CC), cingulum bundle (CG), corticospinal tract (CST), fornix (FX), inferior fronto-occipital fasciculus (IFOF), inferior longitudinal fasciculus (ILF), superior cerebral peduncle (SCP), three segments of the superior longitudinal fasciculus (SLF I, II, and III), and uncinate fasciculus (UF). With the exception of the corpus callosum, all tracts were bilateral (one left hemisphere tract and one right hemisphere tract).

Delineated fibre tracts were then converted into fixel masks using the ‘tck2fixel’ command in MRtrix3 ^23^, and subsequently thresholding this fixel image to create a binary tract fixel mask. The mean FDC value across all fixels within each tract of interest was computed for each participant. Given that DWI data from patients included in this study were acquired from two different scanners, we firstly performed tract data harmonization using ComBat^18^. Following this, assumptions of normality were tested for each tract across the healthy control cohort, after which rank-based inverse normal transformations (INTs) were applied.

The mean and standard deviation (SD) of tract FDC was computed across the healthy control cohort for each white matter tract. Following this, Z-scores were calculated for individual patients with epilepsy when compared to the healthy control cohort. Spider plots were used to display tract Z-scores for individual patients across all tracts. The pipeline for individual tract-based analysis is shown in Figure 1.

**Figure 1:**
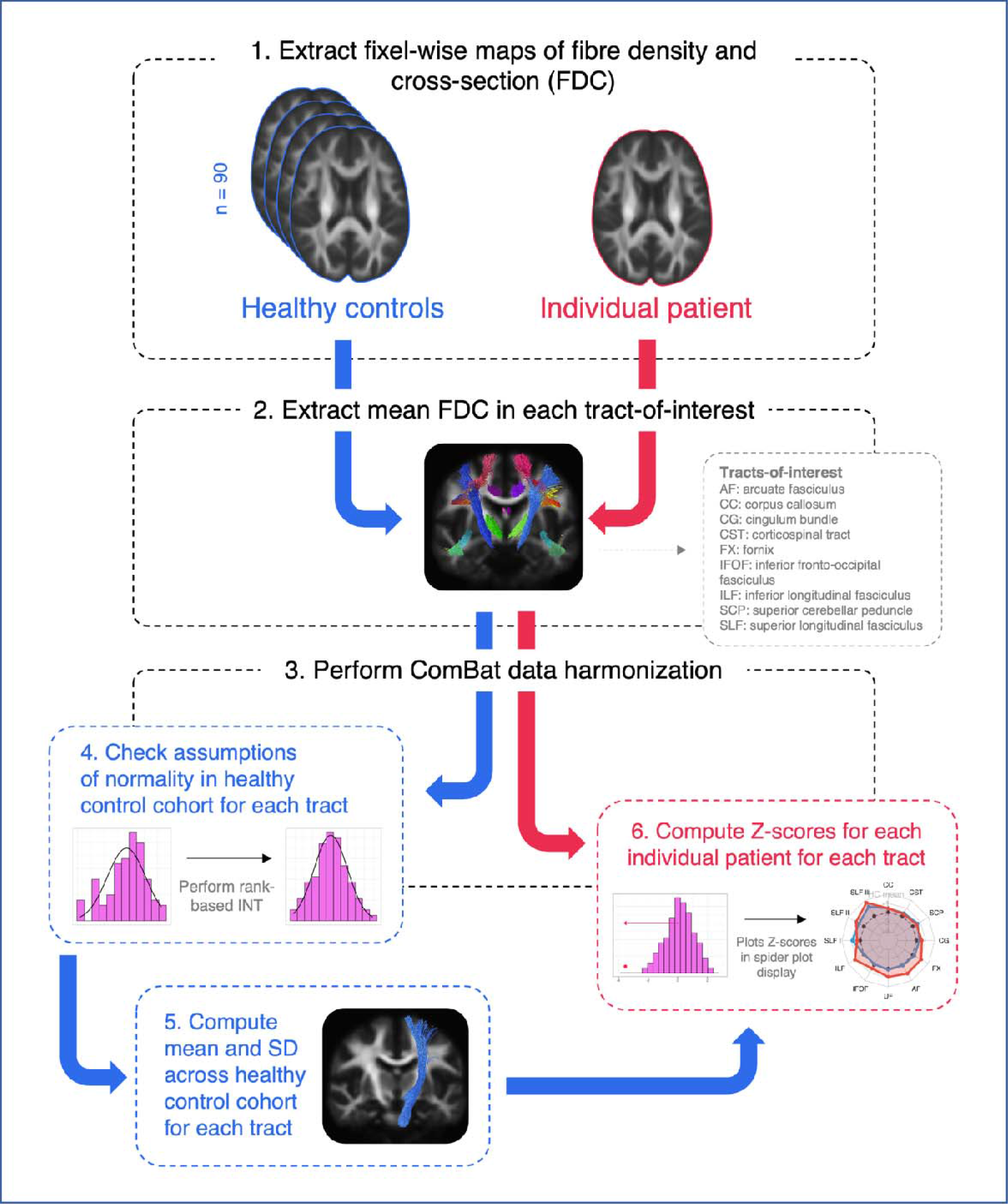
Methodology for individual tract-based FDC analysis. 1. A measure of fibre density and cross-section (FDC) is quantified at each white matter fixel in template space, for all healthy control participants and individuals with epilepsy. 2. Mean FDC is computed within select tracts-of-interest. 3. ComBat data harmonization is performed on the tract data to adjust for any scanner effects. 4. Assumptions of normality are tested across the control cohort. 5. The mean and standard deviation (SD) are computed across the healthy control cohort for each tract. 6. Tract-wise Z-scores are computed for each patient.

A summary laterality index (LI) was computed using the resepective sums of all Z-scores across all tracts in each hemisphere: LI = L_sum_ – R_sum_ / L_sum_ + R_sum_.

All tract-level analysis was performed in R statistical software (version 3.6.3). Packages utilised include the ‘neuroCombat’ package for tract data harmonization, ‘RNOmni’ package^40^ to perform rank-based INTs, and ‘fmsb’ package^41^ for generation of spider plots. The code to perform analysis and generate plots is available in R, along with tract data from this study (https://github.com/remikamito/spidey).

### Fixel-level analysis

Subject-specific deviations from the healthy control population were also explored at the level of each individual white matter fixel. Here, a similar pipeline to the tract-based approach was implemented, whereby the healthy population FDC mean and standard deviation were computed at each white matter fixel. Z-scores were then calculated at each template fixel for each epilepsy patient to create a Z-map of fixel-level deviations compared to the control cohort.

## Results

The results of the individualised fixel analysis framework are illustrated with 10 case studies. Table 2 summarises the key findings for each patient, including a summary laterality index for tract-based results, and white matter regions that exhibited decreased FDC in the whole-brain fixel-level maps. As expected, individual patients with epilepsy exhibited highly variable patterns of white matter abnormality compared to the healthy control cohort, both at the tract-level and fixel-level. Figure 2 is a demonstration of the spider plot display that is used to present patient tract-level results in Figures 3-5 (data from Patient 1).

**Table 2:** Key tract-level and fixel-level findings for each patient.

| Patient | Tract-level findings |  |  | Key fixel-level findings |
| --- | --- | --- | --- | --- |
|  | Tract Z-score laterality index<sup>1</sup> | Most affected tract | Smallest Z-score |  |
| **1** | 0.1538 | Left IFOF | -2.13 | Relatively extensive white matter abnormalities in fixels within temporal lobe (L>R) and association fibre pathways connecting into L inferior frontal lobe, along with thalamocortical pathways bilaterally |
| **2** | -0.136 | Right UF | -2.73 | Some diffuse white matter abnormalities bilaterally, but with greatest cluster of abnormality within right temporal lobe and connecting into thalamic structures |
| **3** | -0.258 | Left UF | -1.00 | Small clusters of diffuse abnormality, including small areas within right temporal lobe extending into inferior frontal lobe, along with left inferior frontal cluster |
| **4** | 0.063 | Left cingulum | -1.22 | Some diffuse white matter abnormalities in temporal lobes bilaterally (L>R), along with bilateral thalami and splenium of corpus callosum |
| **5** | 0.028 | Left SCP | -2.74 | Focal, bilateral white matter abnormality in cerebellar white matter |
| **6** | 0.018 | Right CST | -2.79 | Extensive and bilateral white matter abnormality throughout most brain white matter |
| **7** | 0.393 | Left FX | -2.26 | White matter abnormalities in left cingulum bundle, splenium of corpus callosum and bilateral fornices, and surrounding posterior horn of lateral ventricles (L>R), and left medial temporal lobe |
| **8** | -1.594 | Right AF | -2.04 | Right hemisphere white matter abnormality within temporal lobe, white matter surrounding posterior horn of lateral ventricle and extending into parietal white matter superiorly |
| **9** | -0.163 | Right CST | -2.19 | Major focus of white matter abnormality in right inferior parietal/posterior frontal white matter surrounding post-central sulcus |
| **10** | 0.121 | Left SCP | -0.83 | No sizeable clusters of white matter abnormality observed, although some diffuse subtle abnormalities observed in right central sulcus and right superior cerebellar white matter |

<sup>1</sup>Positive values indicate L>R abnormality; Negative indicates R>L abnormality. Tract laterality is computed across all included tracts in each hemisphere.

**Figure 2:**
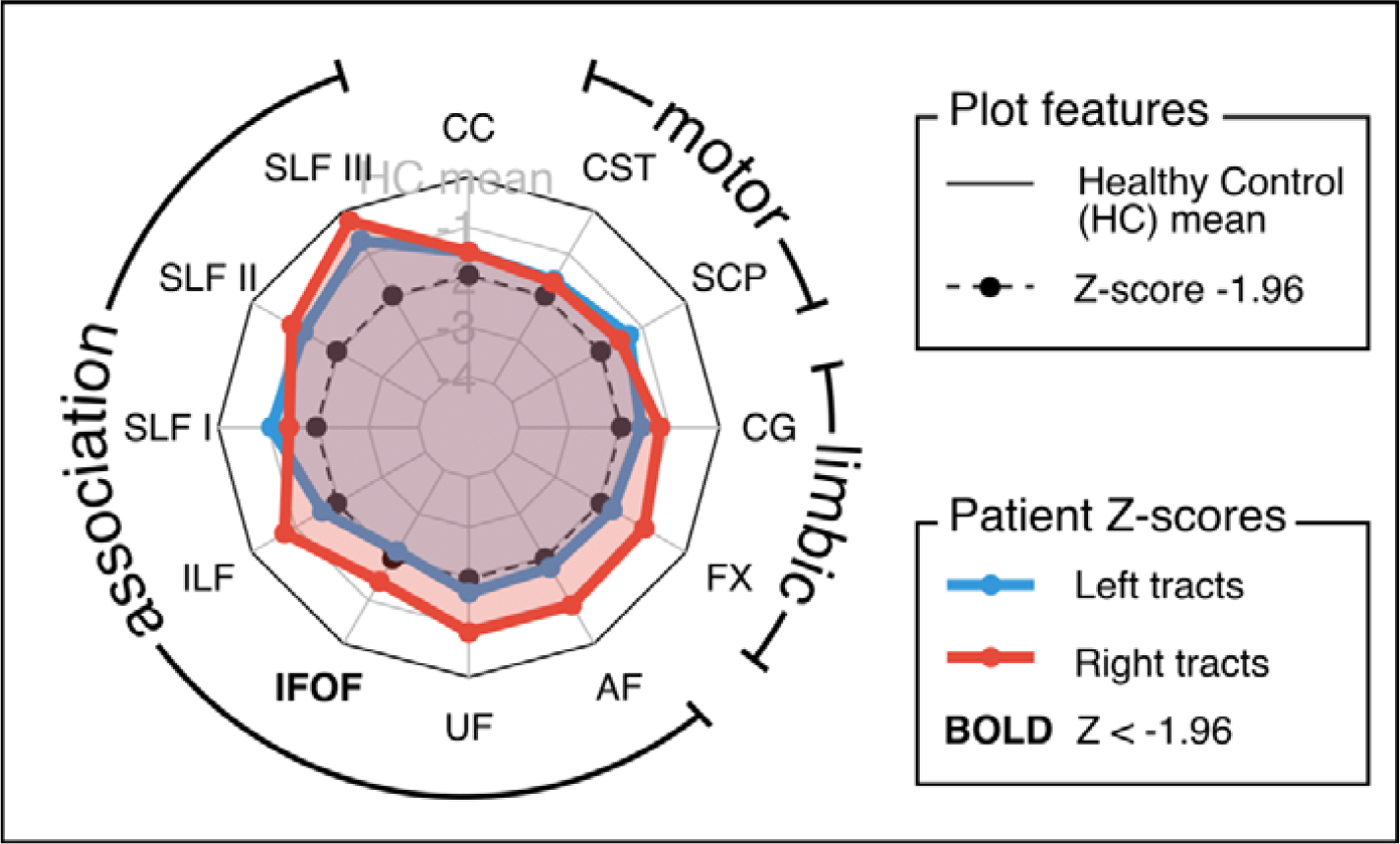
Exemplar spider plot display for tract Z-scores in an individual patient. Z-scores are plotted for each of the included fibre tracts in a spider plot display. The inner dotted line reflects a Z-score of -1.96, and outer solid line reflects the healthy control mean. Z-scores for left hemisphere tracts are shown in blue, while Z-scores for right hemisphere tracts are shown in red. The corpus callosum is included at the top centre, and the Z-score for this tract is included in both left and right hemisphere spider plots. Tracts with a Z-score of less than -1.96 in either hemisphere are labelled in bold.

**Figure 3:**
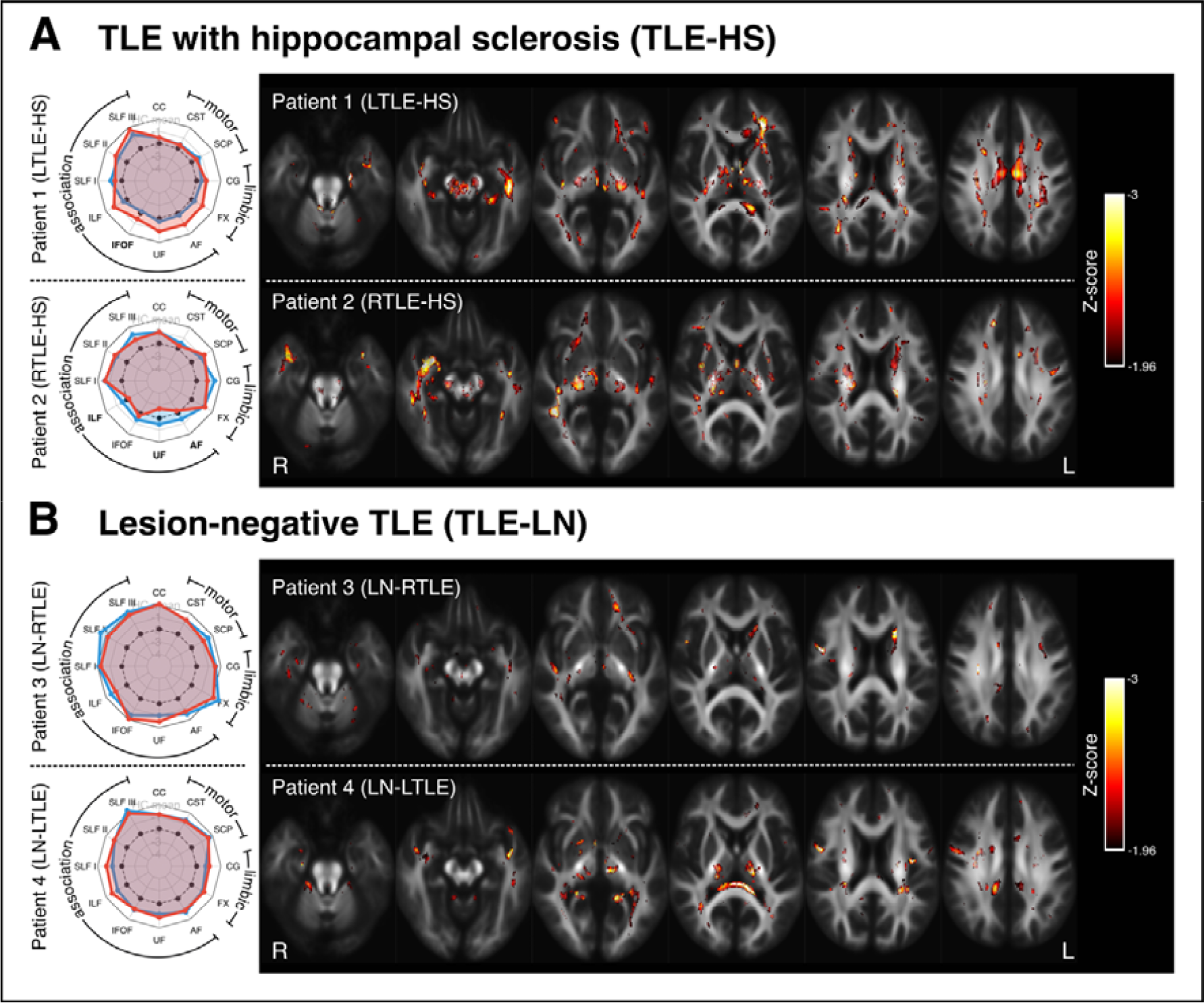
Tract- and fixel-level results for four patients with temporal lobe epilepsy (TLE). A: Two patients with hippocampal sclerosis (HS): Patient 1 is an individual who has left TLE with hippocampal sclerosis (L TLE-HS); Patient 2 is an individual with right TLE with HS (R TLE-HS). B: Two patients with lesion-negative TLE: Patient 3 is an individual with MRI-negative or lesion-negative right TLE (LN-RTLE); and Patient 4 is an individual with lesion-negative left TLE (LN-LTLE). In all cases, spider plots on the left show Z-scores for fibre density and cross-section (FDC) measures in each tract-of-interest (see Figure 2 for interpretation). Fixel-level Z-score maps are shown on the right, with fixels exhibiting a Z-score of -1.96 or greater. Axial slices are placed at 10 mm increments.

### Patients 1-4: Temporal Lobe Epilepsy

Figure 3 shows the tract- and fixel-level results for the four patients with temporal lobe epilepsy (Patients 1-4). Patient 1 is an individual with severe left HS and recurrent focal dyscognitive seizures. Patient 2 is an individual with right TLE due to hippocampal sclerosis. Patient 3 is an individual with MRI lesion-negative right TLE (LN-RTLE). Patient 4 is an individual with lesion-negative left TLE (LN-LTLE).

In patients with TLE and HS (Patients 1 and 2), tract-level spider plots demonstrate predominantly ipsilateral atrophy of tracts related to the temporal lobe (Table 2), with greatest FDC reductions in the UF, AF, ILF, and IFOF. Fixel-level abnormalities were greatest in the affected medial temporal lobe for both patients (left for Patient 1, right for Patient 2).

The patients with lesion-negative TLE (Patients 3 and 4) had milder but diffuse bilateral tract changes, with slightly greater tract abnormality ipsilateral to the epileptic side. Fixel-level findings did not indicate any striking focal abnormalities in these two individuals.

### Patients 5 and 6: Progressive Myoclonus Epilepsy and Dravet Syndrome

Figure 4 shows tract and fixel-level results for two examples of patients with specific epilepsy syndromes: Progressive Myoclonus Epilepsy (PME) and Dravet Syndrome.

**Figure 4:**
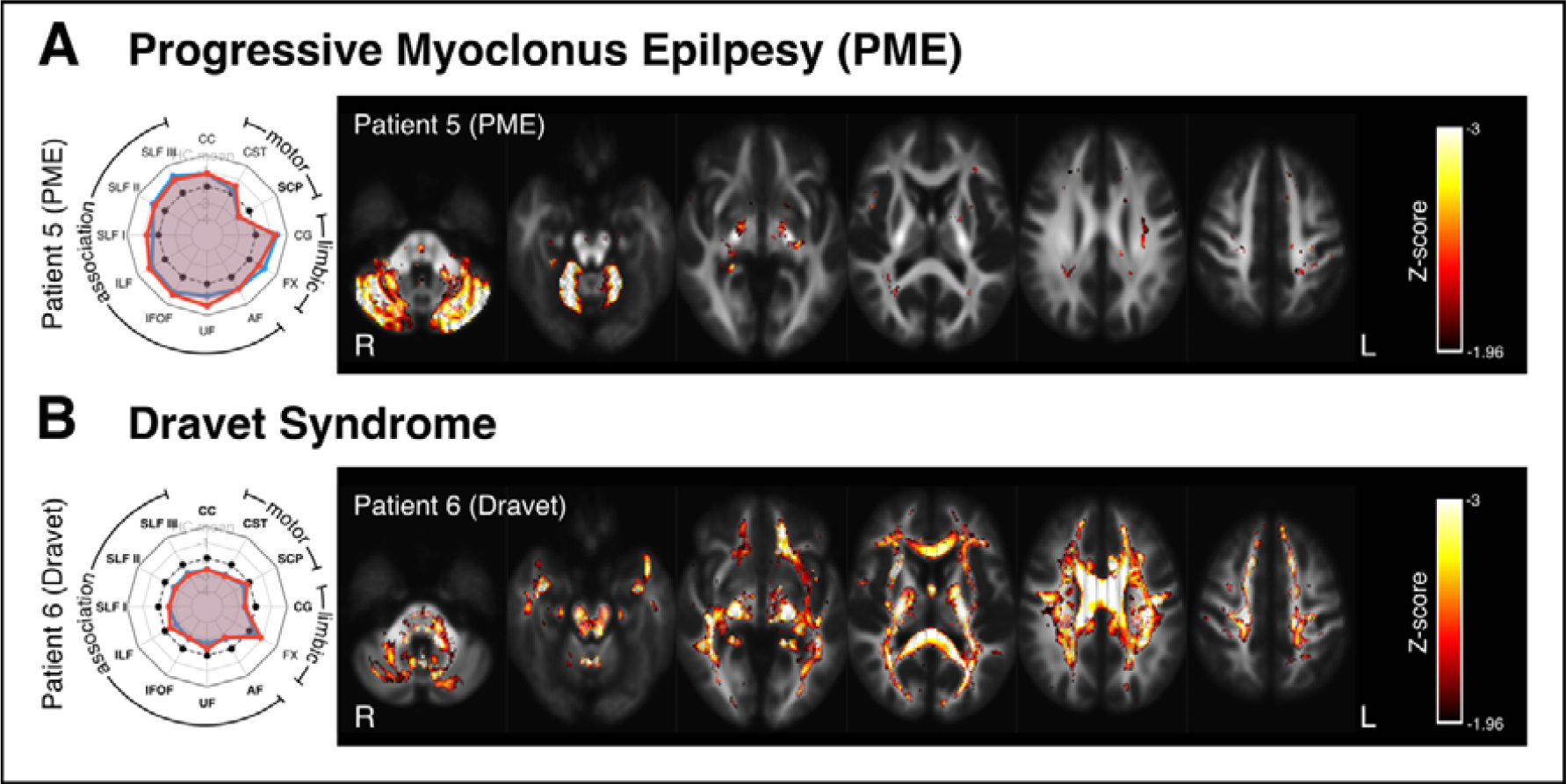
Results in Syndromic Epilepsies. A: Patient 5 is an individual with Progressive Myoclonus Epilepsy. B: Patient 6 is an individual with Dravet syndrome. As with Figure 3, spider plots are displayed on the left, showing tract-based Z-scores, while fixel Z-maps are displayed on the right. Axial slices are placed at 15mm increments.

Patient 5 is an individual with PME with reasonable cognitive function. PME is commonly associated with cerebellar abnormalities, and the tract-level analysis revealed striking bilateral abnormality within the superior cerebellar peduncles (SCP), with minimal deviation from the norm in most other fibre tracts. The fixel-level Z-score map similarly exhibited focal abnormality within cerebellar white matter.

Patient 6 is an individual with severe Dravet syndrome and intellectual disability. Both tract-level and fixel-level FDC measures exhibited widespread abnormality across most white matter fibre tracts.

### Patients 7-10: Malformations of cortical development

Figure 5 shows both tract-level spider plots and fixel-level findings in patients with malformations of cortical development (Patients 7-10). Patient 7 is an individual with focal dyscognitive seizures due to periventricular nodular heterotopia (PVNH) surrounding the left lateral ventricles. Patient 8 is an individual with focal epilepsy due to periventricular nodular heterotopia (PVNH) and polymicrogyria within the right temporal lobe. Patient 9 is an individual with a small bottom-of-sulcus dysplasia (BOSD) in the right precentral sulcus, and Patient 10 is an individual with a small bottom-of-sulcus dysplasia in the right postcentral sulcus.

**Figure 5:**
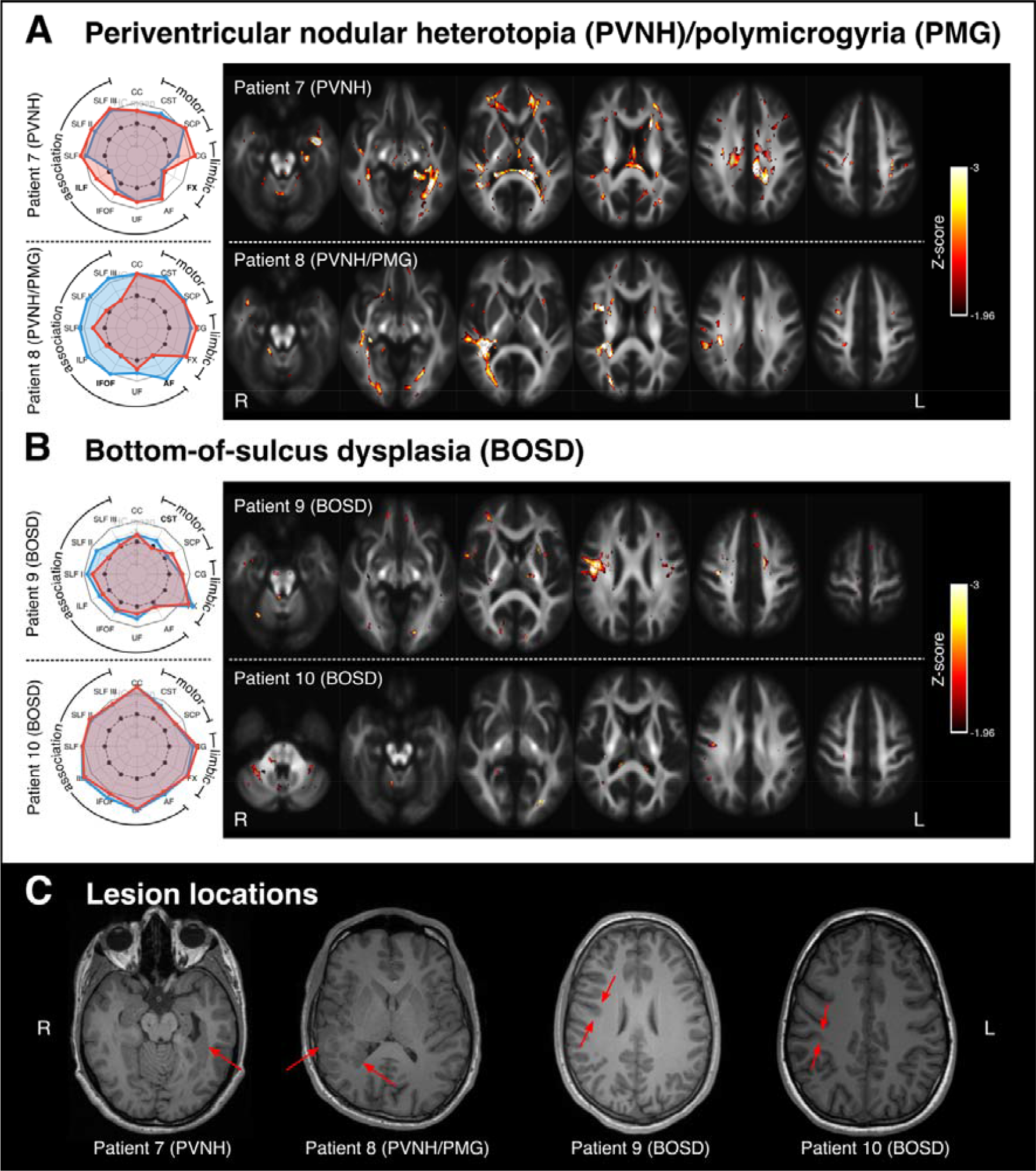
Results in lesional epilepsies. Panel A shows individual-level results for two patients with periventricular nodular heterotopia (PVNH), one with additional polymicrogyria (PMG). In both cases, lesions were unilateral, with Patient 7 exhibiting left-sided PVNH, and Patient 8 exhibiting right-sided PVNH and PMG. Panel B shows individual-level results for two patients with bottom-of-sulcus dysplasia (BOSD): Patient 9 had a small BOSD in the right precentral sulcus; Patient 10 had a BOSD in the right postcentral sulcus. Panel C shows the lesion locations for all patients (Patients 7-10) on a single axial slice from a T1-weighted image in each subject’s own space.

In all patients, the tract laterality index indicated greater tract abnormality on the side ipsilateral to the respective lesion(s). Tract-level findings also appear to reflect the clinical or cognitive profiles in these individuals: for example, in Patient 8, left hemisphere tracts appear normal while there is notable right hemisphere white matter abnormality, in keeping with the cognitive profile for this individual, who had normal intellect but some non-verbal memory deficits.

In most cases, either the most affected tract or the pattern of tract-level changes provided some indication of the spatial location of the structural epileptogenic abnormality; however, tract-level changes were of insufficient granularity to identify the focal lesion, particularly in the two cases with bottom-of-sulcus dysplasia. Importantly, fixel-level Z-maps provided finer grain detail, exhibiting patterns of white matter abnormality that were concordant with the epileptogenic lesions. In the two patients with highly focal BOSD, although Patient 9 exhibited a striking fixel-level finding that was spatially concordant with the epileptogenic lesion (right precentral sulcus BOSD), Patient 10 exhibited only subtle white matter changes, with small foci of decreased FDC and only a small cluster of potentially concordant change in the right central region near the lesion (right postcentral BOSD).

## Discussion

We present an advanced diffusion MRI approach quantifying white matter abnormalities in individual epilepsy patients, by adapting the fixel-based analysis (FBA) framework to individualised assessment. This approach provides proof-of-principle evidence of the potential clinical use of this advanced diffusion MRI technique in epilepsy. In this case series, we show individual-level findings of tract- and fixel-specific change in patients with varying types and causes of epilepsy (TLE, PME, Dravet syndrome and MCDs). We demonstrate that patients with epilepsy exhibit variable patterns of white matter abnormalities and that these white matter changes are concordant with the clinical profile of these patients.

### Individualised white matter abnormalities reflect syndrome-specific patterns

In this study, we assessed fixel-based metrics in 10 patients with varying epilepsy causes and syndromes. This enabled identification of individualised differences within white matter fibre tracts when compared to a control cohort. The patterns of white matter abnormality in these patients appeared concordant with the specific epilepsy syndrome or cause in each individual. However, patient-level variability was also observed, highlighting potentially heterogeneous white matter profiles across individuals, particularly in those with spatially distributed focal lesions.

For the patients with temporal lobe epilepsy, variable patterns of white matter abnormality were observed. Although white matter abnormalities appeared particularly marked within the temporal lobe ipsilateral to the lesion in the two patients with hippocampal sclerosis, they also extended beyond the temporal lobes. The tract-specific decreases in fibre density and cross-section were more marked in the two patients with hippocampal sclerosis (Patients 1 and 2), than in the two lesion-negative TLE patients. Indeed, the two lesion-negative TLE patients exhibited relatively minimal white matter abnormality. This is consistent with group-level findings, which have demonstrated more extensive patterns of abnormality in patients with hippocampal sclerosis than in MRI-negative TLE, both within white matter^2,4,5^ and in the cortex^42,43^. However, these differences have been suggested to be influenced by clinical characteristics such as disease duration^2^. Of note, both MRI-negative TLE patients in our study had later age of onset and shorter disease duration than the two TLE-HS patients, and the difference in extent of white matter abnormalities may reflect these clinical characteristics.

In contrast to the patients with TLE, the two patients with specific epilepsy syndromes associated with generalised seizure types exhibited much more bilateral patterns of white matter abnormality. The patient with progressive myoclonus epilepsy (PME) exhibited a highly cerebellar signature in their pattern of white matter abnormality. Patients with PME exhibit progressive neurological dysfunction, including cerebellar ataxia, and the causes of PME are often associated with cerebellar pathology that is absent in the cerebrum^44^. The pattern of tract-specific white matter abnormality in the PME patient included in this study (Patient 5) suggested a marked cerebellar focus on this individual’s pathology, which is concordant with the clinical signature of PME. Although this individual did not have the more common form of Unverricht-Lundborg Disease, a group study in ULD using DTI has similarly demonstrated cerebellar, subcortical, and thalamocortical white matter changes^45^.

Dravet Syndrome is a severe drug-resistant epilepsy syndrome characterised by generalised tonic-clonic seizures, often with intellectual disability. The patient with Dravet syndrome (Patient 6) exhibited widespread abnormality in white matter tract FDC, with virtually all tracts of interest exhibiting substantial decreases in FDC, and much of the brain’s white matter being implicated at the fixel-level. This finding is in line with the marked white matter changes demonstrated at a group level using fixel-based analysis^46,47^, which may result from abnormal axonal growth in Dravet syndrome, as shown in animal models^48^.

Unlike patients with TLE or specific generalised epilepsy syndromes, where the observed white matter alterations at the individual level appeared concordant with previous group-level findings, people with malformations of cortical development exhibited highly patient-specific changes. In three of these four individuals (Patients 7 – 9), the pattern of white matter abnormality was spatially concordant with the identified seizure-causing structural abnormality. Patients 7 and 8, who had unilateral periventricular nodular heterotopia (with additional polymicrogyria in Patient 8), exhibited white matter abnormalities in a highly lateralised manner, with the most notable abnormalities in the white matter proximal to the heterotopic lesions. Patient 9 exhibited focal abnormality in the right precentral region, spatially concordant with the epileptogenic BOSD in this individual, while Patient 10, who had somewhat more subtle BOSD in a spatially proximal area (right postcentral region), did not show any notable white matter changes.

Group studies comparing patients with malformations of cortical development to healthy control cohorts have typically shown white matter changes that extend beyond the MRI-visible lesions^4,6,49–54^. In the case of BOSD patients, we recently showed that despite spatial heterogeneity in the location of the epileptogenic lesions, patients exhibited a typical pattern of bilateral structural network abnormality^6^. These bilaterally distributed white matter changes have been suggested to reflect the secondary effect of epilepsy on the brain^6,52^, rather than primary pathology in these patients, given the heterogeneous distribution of these lesions. In group-level analyses, we are likely sensitive to such seizure-affected white matter changes, which may be common to patients within an otherwise quite heterogeneous group, while being largely desensitised to subject-specific abnormalities. On the other hand, individual-level abnormalities are likely to be more marked in the area immediately proximal to these epileptogenic lesions, and may be more clinically meaningful than group-level findings.

### Clinical significance of individual tract-specific analysis

An important consideration in MRI studies in epilepsy, particularly in patient cohorts with spatially varied pathology, is the heterogeneity within patient groups. Group-based studies inherently compare group means, which enables identification of co-localised effects between subject groups, but disregard individual-level differences outside these co-localised regions as noise. Neuroimaging studies over the past decade, including large collaborative studies^2,42^, have predominantly focused on group-level analyses that demonstrate common brain changes in patients with varying types and causes of epilepsy.

There is growing interest in moving from group-level studies to individualised neuroimaging analysis and prediction in epilepsy^55^. This is particularly pertinent for precision medicine in epilepsy, where neuroimaging plays a key role in diagnosis and clinical management. To meet this need, neuroimaging techniques must offer clinically valuable insights at an individual patient level. This study examined whether white matter abnormalities in individual patients with varying types and causes of epilepsy offer clinically insightful information, and translate an advanced diffusion MRI tool into the individual-patient level.

In light of the observed abnormalities in the individual patients included in this study, we believe the clinical significance of this individual-level fixel-based analysis is two-fold. Firstly, tract-specific white matter changes may provide insight into the underlying epilepsy type or syndrome, and we may be able to detect patterns of white matter disruption that are diagnostically useful. Secondly, individual-level Z-score maps of fibre tract-specific abnormality may help to localise focal epileptogenic abnormalities, particularly in cases where these abnormalities are highly subtle (e.g., in bottom-of-sulcus dysplasia).

We note that our study is not the first to investigate white matter abnormalities in individual patients with diffusion MRI. As with group-level analyses, studies examining white matter abnormalities in individual patients with epilepsy have typically adopted the diffusion tensor model, demonstrating subject-specific microstructural abnormalities ^49,56^. The voxel-based or region-of-interest-based findings from DTI studies have been interpreted to identify affected fibre structures in epilepsy patients. However, DTI-based results are inherently voxel-averaged or voxel-aggregated, rendering them potentially sensitive, though not specific to underlying fibre structures^15,57^. Although advanced diffusion MRI methods have similarly been applied to probe microstructural changes^11,58^, these studies have also depended on measures that are quantified on a per-voxel basis.

Importantly, by implementing a fixel-based analysis approach, this work moves beyond voxel-averaged measures to a fibre-tract-specific model^20,59^. This approach enables more comprehensive insight into white matter changes within specific fibre populations, as has been demonstrated extensively in many group-level analyses^60^. By utilising a fixel-based approach, reported changes to the derived metrics (namely, fibre density and cross-section) can be ascribed to specific fibre tracts. In this work, we demonstrate that this advanced diffusion MRI approach can be extended to applicability at an individual patient level, and in doing so, can offer important clinical insight into the specific fibre tracts that may be affected in individuals.

### Limitations and future directions

Despite the promise of this individualised approach, there are limitations to this work. This study should be considered proof-of-principle, underscoring the importance of developing a methodological approach for detecting individual tract-specific changes. As such, we included a set of epilepsy patients (n=10) with variable causes of epilepsy to gauge the types of patients in whom these individualised analyses may be helpful in future. We also selected patients from retrospective cohorts who exhibited well-characterised clinical signs typical of their epilepsy diagnosis. It is important to note that, unlike group-level analyses, improvements in statistical power cannot be achieved in this approach by including a greater number of patients. However, future studies with larger cohorts will provide a clearer picture of individual changes in white matter in the epilepsies, both within and across diagnoses. To this end, the code and data used to generate individualised tract-based changes are included in this work. Future work should systematically test individual-level tract changes in a large cohort of epilepsy patients, including those with complex or atypical aetiology, to determine if the clinical utility of this technique.

In this work, we focused on the spatial localisation of any white matter changes observed, rather than on quantifying the significance of any affect, and therefore did not perform stringent statistical corrections for multiple comparisons. It is essential to note that clinical tools at the individual level should focus on quantifying effect size over statistical significance. Setting somewhat arbitrary thresholds for statistical significance (e.g., by performing false discovery rate or family-wise error corrections as has been the case in other individualised approaches^61^) runs the risk of disregarding potentially important clinical information; however, the converse of excess false positives may complicate interpretation for clinicians. To this end, approaches that capture variation across multiple clinical and demographic parameters within large control cohorts may be valuable, as they will likely help to identify key deviations that are of biological or clinical importance.

One approach to circumvent the challenges of classical statistical inference methods in individual patient analysis is to adopt machine learning approaches. In particular, normative modelling approaches that incorporate machine learning models have gained substantial momentum in the last few years, as a means to probe single subject-level deviations against a large normative sample^62,63^. Recent work in other neurological conditions and disorders, including psychiatric disorders^63,64^ and dementia^65^, has demonstrated that individual-level brain structural abnormalities can be identified using such normative approaches. To a limited extent, microstructural abnormalities have been demonstrated in patients with focal cortical dysplasia^66^ using normative diffusion MRI-based models, highlighting the approach’s potential in epilepsy. Normative modelling approaches can be applied both independently to select measures (e.g., independent Z-scores per tract), or to extract multivariate measures of deviation (e.g., spatial Z-maps across brain measures), and by implementing machine learning algorithms that are trained on normative features, these approaches are likely to be more sensitive to than classical univariate frameworks ^66^. However, given that these machine learning approaches rely on computationally expensive training utilising big datasets, we believe that proof-of-principal studies such as this will play an important role in firstly validating the potential clinical impact of a given approach prior to generating large-scale models, in order to minimise wasted resources^67^.

## Conclusion

For neuroimaging studies to be implemented in clinical use, it is crucial to quantify brain abnormalities in individual patients, rather than detect group-level differences between patient and control populations. Our study provides proof-of-principle evidence that by using advanced diffusion MRI analysis, we can identify tract-specific white matter abnormalities in individual patients with various types of epilepsy. Our findings suggest that individual white-matter abnormalities could provide clinically useful information, as the observations appear either concordant with the epilepsy syndromes, or spatially concordant with structural abnormalities. We believe these findings are a step towards moving imaging techniques from group-level studies to clinical relevance, supporting precision medicine for the individual patient in future. We have released codes for creating individualised plots for Z-statistics to further encourage other research groups to assess the clinical potential of this approach.

## Data Availability

All data produced in the present study are available upon reasonable request to the authors. The code to perform analysis and generate plots is available online, along with tract data from this study.

https://github.com/remikamito/spidey

## Acknowledgements

The authors would like to thank all the participants and their families for their involvement in this study. The Florey Institute of Neuroscience and Mental Health acknowledges the strong support from the Victorian Government and in particular the funding from the Operational Infrastructure Support Grant. The authors acknowledge the facilities and scientific and technical assistance of the National Imaging Facility, a National Collaborative Research Infrastructure Strategy (NCRIS) capability, at the Florey Institute of Neuroscience and Mental Health.

